# Dolutegravir-related hyperglycemia among children and adolescents <18 years in Northern and Eastern Uganda: A cross-sectional study

**DOI:** 10.1101/2023.12.05.23299497

**Authors:** Fridah Akello, Damalie Nalwanga, Victor Musiime, Sarah Kiguli

## Abstract

**Background:** The World Health Organization (WHO) recommended Dolutegravir (DTG) as the preferred anchor drug for first and second line Anti-Retroviral Treatment (ART) regimen. Case reports and studies have demonstrated new onset hyperglycemia among adults initiated or transitioned to DTG. There is paucity of data on DTG-related hyperglycemia among children and adolescents and subsequently, glycemic screening is not routinely conducted for those using DTG. We determined the prevalence and factors associated with DTG-related hyperglycemia in this age category.

**Methods:** This cross-sectional study was conducted from 28^th^ October 2022 to 11^th^ September 2023 at Soroti and Lira Regional Referral Hospitals. 251 children and adolescents under 18 years, on a DTG based ART regimen, were consecutively enrolled and data analyzed for 240 participants. Socio-demographic, anthropometric measurements, clinical, and laboratory variables were collected using a questionnaire. Random blood glucose testing, urine dipstick and HbA1c analyses were conducted. Data was entered into epiData version 4.6, cleaned, and analyzed using STATA version 17.0. Logistic regression was used to determine the relationship between outcome and predictor variables. A p-value < 0.05 was considered statistically significant at multivariate analysis.

**Results:** Of the 240 study participants, 55.4 %(n=133) were female and 71.7 %(n =172) were over 10 years. The prevalence of DTG-related-hyperglycemia was 20% (n=49) and 95.9% (n=47) met the criteria for pre-diabetes. Health education talks on Adverse events related to Dolutegravir was protective for hyperglycemia **(aOR 0.4, 95% CI 0.2-0.9), p = 0.035.**

**Conclusion:** The prevalence of DTG-related-hyperglycemia was high, occurring in every 2 in 10 children and adolescents. Health education talks on adverse events associated with DTG use were found to be protective against hyperglycemia. Routine glycemic screening and monitoring therefore should be emphasized. Additionally, a strategy for health education talks focusing on lifestyle modifications to reduce the occurrence of hyperglycemia should be developed.

## Introduction

The world Health Organization and Ministry of Health recommended Dolutegravir (DTG), an Integrase Strand Transfer Inhibitor (INSTI) as the preferred anchor drug for first and second line HIV treatment options (1). This was supported by study results from the ODYSSEY (Once-daily Dolutegravir based ART in Young people versus Standard Therapy) and Penta-17 trials that showed Dolutegravir to be superior to the current standard of care ART regimen (predominantly a Protease Inhibitor based-ART) in children and adolescents (2).

Subsequently, all eligible persons living with HIV are being initiated or optimized to a DTG containing regimen however, a number of case reports and studies among adult patients document symptomatic hyperglycemia associated with initiation on DTG (3–5). Hyperglycemia is defined as a fasting blood glucose level of >7.0 mmol/L or a random plasma glucose greater than 11.1 mmol/L and or an HBA1C level >6.5%(6). The prediabetes state defined by the American Diabetic association as a state of intermediate hyperglycemia based on blood glucose levels or estimates of glycated haemoglobin levels is also considered hyperglycemia.

DTG is postulated to cause hyperglycemia through chelation of magnesium to prevent HIV from integrating into host DNA(7,8). Because magnesium serves as a cofactor in post-receptor insulin action, Magnesium chelation may cause disorders of glucose metabolism (9,10). In addition, concomitant HIV infection is associated with insulin resistance resulting from immune activation and chronic inflammation thus worsening the hyperglycemia (11).

Despite these isolated study findings, there is paucity of data on DTG associated hyperglycemia and even more particularly in the pediatric and adolescent population. Hyperglycemia if undetected or managed late poses severe acute complications including Diabetic Ketoacidosis (DKA) and Hyperosmolar Hyperglycemic State (HHS) (12). Chronic complications are related to the micro vascular changes which predispose to retinopathy, nephropathy and neuropathy resulting from cellular damage inflicted by the hyperglycemia (13).

Ministry of Health (MoH) Uganda HIV treatment guidelines (2022) recommend routine monitoring for therapeutic response and drug related adverse events for all children and adolescents on ART however glycemic monitoring is not specifically mentioned. In contrast however, baseline random blood sugar testing and routine monitoring for adults over 45 years is emphasized in these treatment guidelines. (14). Furthermore, there is scanty data on prevalence and factors associated with DTG-related-hyperglycemia among children and adolescents in Uganda and this study addressed this knowledge gap. Findings from this study form a point of reference for additional studies and guidelines to improve adverse events monitoring for children and adolescents on DTG.

## Methods

### Study site

This was conducted at the HIV clinics of Soroti and Lira Regional Referral Hospitals (RRHs) both tertiary level health care facilities in Eastern and Northern Uganda respectively. Soroti RRH was the primary study site and Lira RRH was included as an additional site having reached saturation in Soroti. Lira RRH was chosen owing to its homogeneity to Soroti in regard to dietary and activity characteristics of the population. Both regions were ravaged by the 5 year LRA war in 2002-2006 and resultantly bore the brunt of the HIV epidemic with relatively high rates of HIV transmission. Each of the hospitals has one of the largest pediatric HIV enrollments in the respective regions and subsequently, a high number of children and adolescents on a DTG based ART regimen. Soroti RRH serves a catchment area with a population of 2.7 million persons. Lira RRH on the other hand offers general and specialized health care services to a population of close to 2.3 million individuals from the 9 districts of Lango sub-region.

### Study population and sample size determination

These were all children and adolescents below 18 years on a DTG containing regimen for at least 6 months attending the paediatric HIV clinics of Soroti and Lira RRHs. The study was conducted from 28^th^ October 2022 to 11^th^ September 2023. The sample size of 240 was determined using the Kish Leslie and Fleiss formulae.

### Study design and data collection procedure

The study used a cross sectional study design. Quantitative data was collected using a questionnaire onto which socio-demographic information, anthropometry and relevant history were captured.

### Inclusion and exclusion criteria

We included all participants below 18 years, on a DTG based regimen for at least 6 months with documented good adherence in the last 4 months and excluded those who had an acute illness, a past history of hyperglycemia or if they declined to consent or assent.

### Study variables

The dependent variable was hyperglycemia defined as a random plasma glucose concentration of ≥11.1 mmol/l (200 mg/dl) or an HbA1c of > 6.5% (15). Patients with prediabetic glycemic levels were also considered to have hyperglycemia. This was determined by a random plasma glucose of 7.8-11.0 mmol/L or an HBA1C of 5.7-6.4%.

Independent variables included patient factors like age, sex, current and pretreatment Body Mass Index (BMI) for age z scores, Pre-existing comorbidities, Age at diagnosis of HIV, Age at ART initiation, Use of other medications, WHO Stage, Duration on ART, Duration on DTG and most recent viral load. Health system factors included Glycemic screening, patient Education on DTG Adverse Events and Glycemic Monitoring

### Data management and analysis

Data was entered into EpiData (v4.6) and exported to STATA version 17 for analysis. Univariate analysis was done for socio-demographic and baseline clinical characteristics and was presented in form of frequencies, proportions and means. A bivariate analysiss was performed to determine the association between the dependent and independent variables. Variables with p values <0.2 were considered for multivariate analysis. A logistic regression was then performed to determine the statistical significance of the association between these variables and hyperglycemia. Factors with P values <0.05 were considered statistically significant.

## Ethical consideration

The study protocol was approved by the School of Medicine Research and Ethics Committee (SOMREC), reference number Mak-SOMREC-2022-405. Administrative clearance was further obtained from the hospital directors and research committees of Soroti and Lira RRHs. Parental consent was obtained for participants below 8 years while assent was obtained for participants 8 years and above and substantiated by a witness within the clinic. All consent was written and documented on a parental consent or assent form as applicable.

## Results

### Participant Demographic and baseline clinical characteristics

Data was analyzed for 240 participants. Mean age was 12.36 ±3.76 years. Females accounted for 55.4% (n=133) and 71.7% (n=172) were over 10 years of age. Median age at HIV diagnosis was 24.06 (IQR, 1.33-46.79) months. Majority of participants, 202 (84.2%), had been on ART for more than 5 years, only 37.9% (n=91) had been on a DTG-containing regimen for more than 2 years while 68.8% (n=165) initiated ART before the age of five years s presented in **Table 1** below.

**Table 1:** Demographic and baseline clinical characteristics of study participants.

| Variable | Total n=240 | Percentage (%) |
| --- | --- | --- |
| **Age (Years)** |  |  |
| 1-4 | 10 | 4.2 |
| 5-10 | 58 | 24.2 |
| >10 | 172 | 71.7 |
| **Sex** |  |  |
| Female | 133 | 55.4 |
| Male | 107 | 44.6 |
| **Age at ART STAT (Years)** |  |  |
| 0-4 | 165 | 68.8 |
| 5-10 | 62 | 25.8 |
| >10 | 13 | 5.4 |
| **Duration on ART(Years)** |  |  |
| <1 | 4 | 1.7 |
| 1-5 | 34 | 14.2 |
| >5 | 202 | 84.2 |
| **Duration on DTG** |  |  |
| <1 | 26 | 10.8 |
| 1-2 | 123 | 51.3 |
| >2 | 91 | 37.9 |
| **Baseline CD4** |  |  |
| <100 cells/mm<sup>3</sup> | 6 | 2.5 |
| 100 <250 cells/mm<sup>3</sup> | 8 | 3.3 |
| 250-500 cells/ mm<sup>3</sup> | 16 | 6.7 |
| >500 cells/ mm<sup>3</sup> | 37 | 15.4 |
| Unknown | 173 | 72.1 |
| **Baseline ART** |  |  |
| NVP based | 128 | 53.3 |
| EFV based | 60 | 25.0 |
| PI based | 40 | 16.7 |
| DTG | 12 | 5.0 |
| **WHO Stage at ART initiation** |  |  |
| Stage I | 177 | 73.8 |
| Stage II | 29 | 12.1 |
| Stage III | 26 | 10.8 |
| Stage IV | 8 | 3.3 |
| **BMI for age z scores at DTG initiation** |  |  |
| Normal | 197 | 82.1 |
| Overweight | 10 | 4.2 |
| Underweight | 11 | 4.6 |
| Not assessed | 22 | 9.1 |
| *Abbreviations: NVP Nevirapine EFV Efavirenz PI Protease Inhibitor* |  |  |

Only 37.9% (n=67) had results for baseline CD4 counts, of which 5.8% (n=14) had a CD4 count below 250 cells/mm3. Approximately 53.3% (n=128) of the study participants initially received Nevirapine as baseline ART, and only 5.0% (n=12) were initiated on a DTG-based regimen.

Significantly, 73.8% (n=177) of the participants were classified as WHO stage I at the initiation of ART, however, upon the most recent WHO clinical staging, this percentage notably increased to 94.6%, highlighting substantial clinical improvement in the study population.

### Recent clinical characteristics of study participants

Majority of clients, 92.1% (n=221), were still on a first-line ART regimen. Specifically, 62.9% (n=151) of participants are on a Tenofovir-based ART drug combination as in **Table 2** below.

**Table 2:** Recent clinical characteristics of study participants.

| Variable | Total n=240 | Percentage (%) |
| --- | --- | --- |
| **Current regimen line** |  |  |
| 1st Line | 221 | 92.1 |
| 2nd Line | 17 | 7.1 |
| 3rd Line | 2 | 0.8 |
| **Current ART Regimen** |  |  |
| ABC based | 80 | 33.3 |
| TDF Based | 151 | 62.9 |
| AZT Based | 8 | 3.3 |
| PI and NRTI based | 1 | 0.4 |
| **Viral Load** |  |  |
| Suppressed | 199 | 82.9 |
| Non suppressed | 38 | 15.8 |
| Not done | 3 | 1.3 |
| **Current BMI for age z scores** |  |  |
| Normal | 205 | 85.4 |
| Overweight | 12 | 5.0 |
| Underweight | 23 | 9.6 |
| **Recent WHO stage** |  |  |
| Stage I | 227 | 94.6 |
| Stage II | 10 | 4.2 |
| Stage III | 2 | 0.8 |
| Stage IV | 1 | 0.4 |

Only 2 (0.9%) participants, have transitioned to a third-line ART regimen. It is noteworthy that only 82.9% of the participants had achieved virological suppression, defined as having a viral load less than 1000 copies/ml.

Additionally, 85.4% (n=205) of the participants had normal body mass index (BMI) for age z-scores and there were notably no significant changes in BMI for age z-scores between the initiation of DTG and recent measurements.

### Prevalence of Dolutegravir-related-hyperglycemia

A participant was considered to have hyperglycemia if either their RBS measurement was greater than 7.8 mmol/L or their HbA1c level was equal to or greater than 5.7%. This inclusive definition also encompassed the prediabetes state. Overall, the findings revealed that 49 (20%) [95% CI 0.1506, 0.2578] study participants had hyperglycemia. Notably, HbA1C levels varied from 3.5% − 6.7%.

Among those diagnosed with hyperglycemia, HbA1c alone was responsible for identifying hyperglycemia in the majority, with 46 (93.9%) participants diagnosed using this measure. Conversely, only a small proportion, 3 (6.1%), were identified as hyperglycemic based on the RBS measurement alone. Only two participants had diabetic range HbA1C levels.

### Patient factors associated with DTG-related-hyperglycemia

At bivariate analysis for the clinical characteristics of study participants, age, duration on DTG and baseline CD4 had P values <0.2 and were considered for the multivariate analysis as shown in **Table 3** below.

**Table 3:** Patient factors associated with DTG-related-hyperglycemia.

| Variable | Hyperglycemia |  | Crude OR (95% CI) | P Value |
| --- | --- | --- | --- | --- |
|  | No<br>n=191 (80%) | Yes<br>n=49 (20%) |  |  |
| **Current regimen line** |  |  |  | 0.071* |
| 1st Line | 172(77.8) | 49(22.2) | 1 |  |
| 2nd Line | 17(100.0) | 0(0.0) | 0.1(0.01, 1.7) | 0.110 |
| 3rd Line | 2(100.0) | 0(0.0) | 0.7(0.03, 14.8) | 0.817 |
| **Current ART Regimen** |  |  |  | 0.172* |
| ABC based | 70(87.5) | 10(12.5) | 1 |  |
| TDF Based | 114(75.5) | 37(24.5) | 2.2(1.04, 4.6) | 0.038* |
| AZT Based | 6(75.0) | 2(25.0) | 2.6(0.5, 12.7) | 0.244 |
| PI and NRTI based | 1(100.0) | 0(0.0) | 2.2(0.1, 58.6) | 0.629 |
| **Viral Load** |  |  |  | 0.308 |
| Suppressed | 155(77.9) | 44(22.1) | 1 |  |
| Non suppressed | 33(86.8) | 5(13.2) | 0.6(0.2, 1.5) | 0.257 |
| Not done | 3(100.0) | 0(0.0) | - |  |
| **Current BMI for age z scores** |  |  |  | 0.791 |
| Normal | 165(80.5) | 40(19.5) | 1 |  |
| Overweight | 8(66.7) | 4(33.3) | 2.2(0.7, 7.1) | 0.205 |
| Underweight | 18(78.3) | 5(21.7) | 1.2(0.4, 3.3) | 0.706 |
| Total | 165(80.5) | 40(19.5) |  |  |
| **Recent WHO stage** |  |  |  | 0.854 |
| Stage 1 | 180(79.3) | 47(20.7) | 1 |  |
| Stage II | 8(80.0) | 2(20.0) | 1.1(0.3, 4.7) | 0.880 |
| Stage IV | 2(100.0) | 0(0.0) | 0.8(0.04, 16.1) | 0.860 |
| Not staged | 1(100.0) | 0(0.0) | 1.3(0.01, 31.6) | 0.885 |
| **Herbal Remedies** |  |  |  | 0.129* |
| No | 174(80.9) | 41(19.1) | 1 |  |
| Yes | 17(68.0) | 8(32.0) | 2.0(0.8, 5.0) | 0.114 |
| **History suggesting hyperglycemia** |  |  |  | 0.001* |
| Polyuria | 3(100.0) | 0(0.0) | - |  |
| Polydipsia | 0(0.0) | 3(100.0) | - |  |
| Polyphagia | 0(0.0) | 1(100.0) | - |  |
| None | 188(80.7) | 45(19.3) | - |  |
| **Family History of HTN** |  |  |  | 0.035* |
| No | 178(81.3) | 41(18.7) | 1 |  |
| Yes | 13(61.9) | 8(38.1) | 2.7(1.1, 6.8) | 0.034 |

At bivariate analysis, current regimen line, current ART regimen, history of use of herbal remedies and family history of hypertension was considered for multivariate analysis.

### Health system factors associated with DTG-related-hyperglycemia

According to the study data, only 45(18.8%) participants had a baseline random blood sugar done at initiation of DTG while 169 (70.4%) had never received any form of glycemic monitoring. The majority of participants, 60.4% had not received a health education talk on DTG related adverse events and only 54.6% were aware of reporting avenues for DTG related adverse events. This is summarized in **Table 4** below.

**Table 4:**
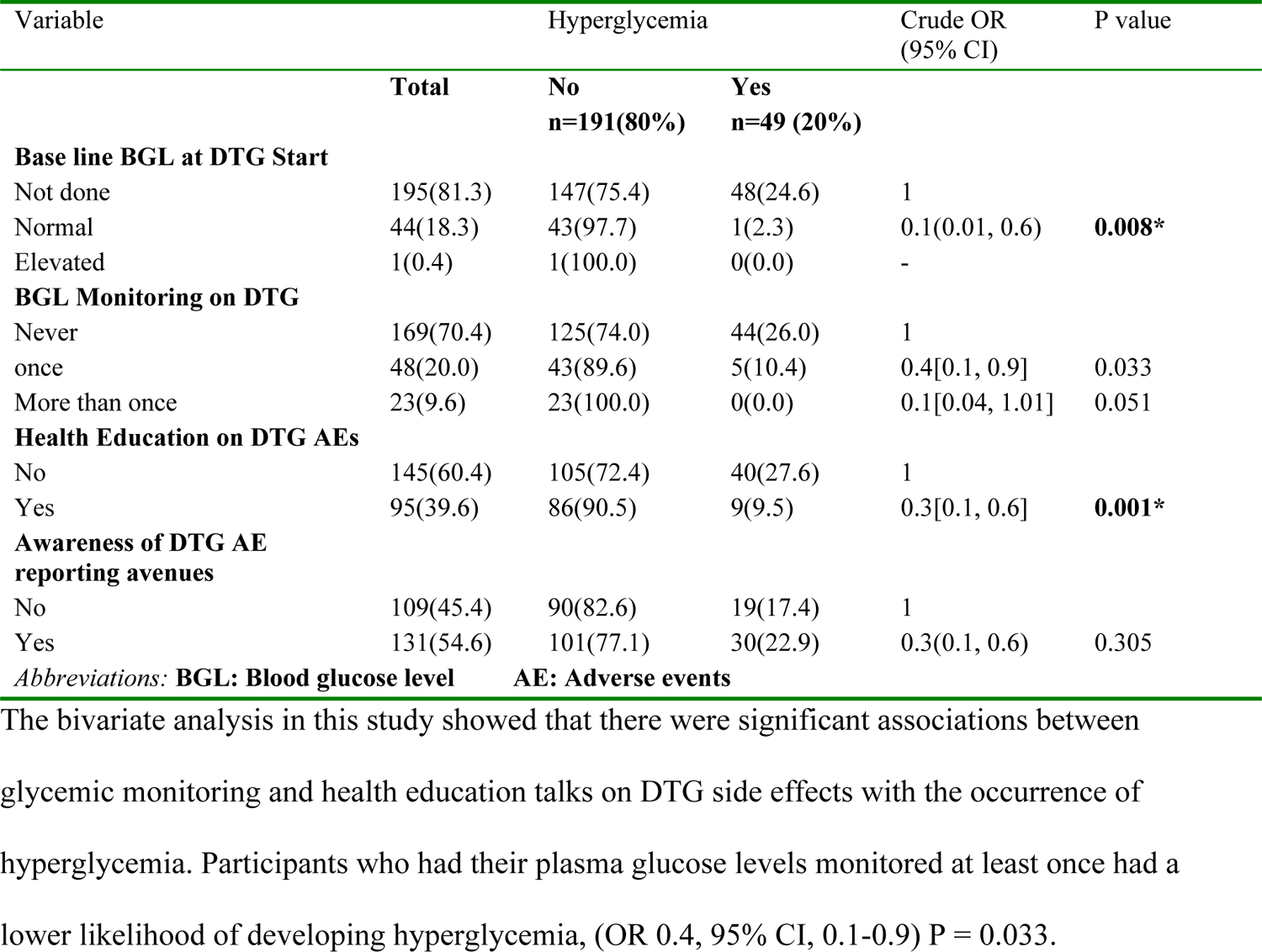
Health System factors associated with DTG-related-hyperglycemia.

The bivariate analysis in this study showed that there were significant associations between glycemic monitoring and health education talks on DTG side effects with the occurrence of hyperglycemia. Participants who had their plasma glucose levels monitored at least once had a lower likelihood of developing hyperglycemia, (OR 0.4, 95% CI, 0.1-0.9) P = 0.033.

Similarly participants who had received a health education talk on the side effects of DTG had a lower likelihood of developing hyperglycemia (OR 0.3, 95% CI, 0.1-0.6), P=0.001.

### Multivariate analysis of factors associated with DTG-related-hyperglycemia

At multivariate analysis, only Health Education on Dolutegravir associated adverse events was found to have a statistically significant association with developing DTG-related hyperglycemia as summarized in **Table 5** below.

**Table 5:** Multivariate analysis of factors associated with Dolutegravir-related-hyperglycemia.

| Variables | COR[95% conf. interval] | P-value | AOR[95% conf. interval] | P-values |
| --- | --- | --- | --- | --- |
| **Duration on DTG[Years]** |  |  |  |  |
| <1 | 1 |  | 1 |  |
| 1-2 | 2.4[0.7, 7.8] | 0.158 | 2.1[0.6, 7.3] | 0.234 |
| >2 | 1.3[0.4, 4.5] | 0.707 | 1.2[0.3, 4.7] | 0.748 |
| **Current ART Regimen** |  |  |  |  |
| ABC based | 1 |  | 1 |  |
| TDF Based | 2.2[1.04, 4.6] | 0.038 | 2.1[0.9, 4.6] | 0.073 |
| AZT Based | 2.6[0.5, 12.7] | 0.244 | 3.4[0.6, 18.7] | 0.153 |
| PI and NRTI based | 2.2[0.1, 58.6] | 0.629 | 5.6[0.2, 170.8] | 0.326 |
| **Family History of HTN** |  |  |  |  |
| No | 1 |  | 1 |  |
| Yes | 2.7[1.1, 6.8] | 0.034 | 1.6[0.6, 4.2] | 0.358 |
| **BGL Monitoring on DTG** |  |  |  |  |
| Never | 1 |  | 1 |  |
| once | 0.4[0.1, 0.9] | 0.033 | 0.4[0.2, 1.2] | 0.101 |
| More than once | 0.1[0.04, 1.01] | 0.051 | 0.1[0.01, 1.7] | 0.113 |
| **Health Education on DTG AEs** |  |  |  |  |
| No | 1 |  | 1 |  |
| Yes | **0.3[0.1, 0.6]** | **0.001** | **0.4[0.2, 0.9]** | **0.035** |

**Abbreviations:** AEs, Adverse events HTN Hypertension ABC Abacavir TDF Tenofovir AZT Zidovudine PI Protease Inhibitor

## Discussion

This cross-sectional study determined the prevalence and factors associated with Dolutegravir-related hyperglycemia among children and adolescents in two regional referral hospitals in Uganda. The study included a total of 240 participants from Soroti RRH and additional participants from Lira Regional Referral Hospital.

Hyperglycemic status was determined based on elevated random blood sugar or glycated hemoglobin levels. The study suggested an association between the use of a Dolutegravir-containing ART regimen and hyperglycemia.

The study findings reveal that among children and adolescents on a DTG-based ART regimen, the prevalence of hyperglycemia was 20%, indicating that approximately two out of every 10 children were affected.

Furthermore, the prevalence of prediabetes and diabetic range hyperglycemia was 17.9% and 2.1% respectively. This prevalence is higher compared to the estimated rates of prediabetes and diabetes in the broader African region, which is reported to be 8.6% and 3.3% respectively (16). The difference in prevalence could be attributed to the specific population studied (children and adolescents on DTG-based ART) who have an additional risk factor of HIV infection, which is known to be associated with an increased risk of hyperglycemia.

Results from a similar study conducted in Botswana by Moverman *et al;*, showed the prevalence of prediabetes and diabetic range hyperglycemia in the general population of persons living with HIV to be 30% and 6% respectively(17). The lower prevalence in our study can be explained by the fact that the former study was conducted in a population of older adults who were over 40 years age and therefore had additional risk factors for hyperglycemia associated with older age.

It is important to highlight that there is currently no published data available on the prevalence of DTG-related hyperglycemia specifically in the child and adolescent age group. Abera *et al;* in 2022 reported a prevalence of 14.7% among adult patients attending a tertiary hospital in Ethiopia. The higher prevalence in our study could be attributed to the inclusion of prediabetic HbA1C levels as part of the criteria for hyperglycemia in the current study, which was not considered in the Ethiopian study. This suggests that prediabetic states may contribute to the higher prevalence observed in the child and adolescent population.

In contrast, Odenyo *et al;* in 2020 reported a prevalence of 55.9% in their study conducted in an adult population in a Kenyan hospital(18). These findings may be attributed to the presence of additional risk factors in the adult age group like insulin resistance, impaired glucose tolerance, or age-related changes in metabolism that predisposes them to an increased risk of developing hyperglycemia.

Related studies include one by Bahamdain *et al;* in Arizona where 7% of adult study participants met the criteria for prediabetes 3-6 months after DTG initiation(19). However, in the current study, due to the cross-sectional study design, the survival time to developing prediabetes could not be determined. In addition, Namara *et al;* in their case control study in Uganda, reported that adults who had been previously exposed to a Dolutegravir-containing regimen had seven times higher odds of developing hyperglycemia compared to those with no history of exposure (20).

Dolutegravir has been postulated to cause hyperglycemia by a mechanism involving chelation of Magnesium which is critical for the action of viral integrase enzyme in the HIV replication cycle. These study findings should therefore be of great concern since current WHO and MoH-Uganda HIV care and treatment guidelines only emphasize DTG glycemic screening and monitoring for mainly adults over 45 years who are perceived to be at risk for hyperglycemia overlooking the potential risk to children and adolescents which is probably based on an assumption that children and adolescents are usually euglycemic. These findings thus suggest that the prevalence of Dolutegravir-related hyperglycemia can vary across different populations and age groups.

At bivariate analysis, a number of factors were found to be associated with hyperglycemia and these included a history of using a TDF based ART regimen, family history of hypertension and history of health education on adverse events associated with DTG use.

Children and adolescents who received health education on the side effects of Dolutegravir were less likely to develop hyperglycemia cOR 0.3 (p = 0.001).Furthermore, at multivariate analysis, after adjusting for other factors, health education talks remained significantly protective against DTG-related hyperglycemia, with (aOR) of 0.4 (p = 0.035). This could be attributed to the fact that currently, facility based education sessions are also geared towards control of Non communicable Diseases (NCDs) with emphasis on enhancement of lifestyle modifications protective for diabetes and hypertension.

There has been a remarkable paradigm shift in the content of health education talks. Early at the start of the HIV epidemic, health education talks were focused on stigma reduction, adherence and disclosure however owing to the success of the ART program, PLHIV are living longer and faced with higher risk for NCDs given the additional risk posed by the HIV infection and the anti HIV drugs. The focus of health education sessions has had to include modalities for screening and prevention of NCDs.

Fitch *et al;* 2006 in their study demonstrated that weekly counseling by a dietician with main emphasis on healthy eating, rather than weight loss, was associated with a significant reduction in blood pressure, waist circumference and HbA1c (21). In addition, endurance and resistance exercises have also been shown to have positive effects on metabolic parameters in HIV infected patients (22) therefore health education messages that highlight such lifestyle modifications are protective for hyperglycemia.

Other studies have shown that effective health education improves knowledge, attitude, and practices, particularly with regard to lifestyle modifications and dietary management, culminating into better glycemic control that can slow down the progression of diabetes and prevent downstream complications (23).

These findings thus highlight the importance of health education in reducing the risk of hyperglycemia in individuals using DTG. The study did not reveal any statistically significant association between hyperglycemia and sex, BMI for age z scores, duration on Dolutegravir, WHO stage, baseline CD4 counts, and the use of herbal drug remedies. These findings differ from previous studies that have reported associations between these factors and Dolutegravir-related hyperglycemia. For example, Odenyo *et al;* found that females were 1.6 times more likely to develop DTG-related hyperglycemia (18), while Miiro *et al;* and Abera *et al;* reported that males were more likely to develop hyperglycemia (24,25). Additionally, Odenyo *et al;* found that overweight patients were 1.7 times more likely to develop hyperglycemia. In terms of BMI for age z scores, the current study’s findings align with studies by Namara *et al;* and the GLUMED study in Kampala, which also did not find a significant association between BMI and hyperglycemia (20,26). The duration on Dolutegravir also did not show a statistically significant association with hyperglycemia in contrast to the findings by Lamorde *et al;* in Uganda (5). Similarly, factors such as WHO stage, baseline CD4 counts, and the use of herbal drug remedies also did not demonstrate a significant association with hyperglycemia in the current study.

### Strength of the study

This is among the few studies in Uganda to investigate the prevalence of DTG-related hyperglycemia among children and adolescents making this study valuable in addressing a critical knowledge gap that highlights the potential risk of hyperglycemia in younger age groups. Additionally, the inclusion of HbA1c as a diagnostic criterion for hyperglycemia adds to the robustness of the study since HbA1c is a reliable indicator of long-term glucose control and provides a comprehensive assessment of glycemic status.

### Study limitations

The study sites were hospitals based in peri-urban settings posing limitations in generalizability to other settings especially the urban rich who are predisposed to cofounders for hyperglycemia like obesity and sedentary lifestyles.

The cross-sectional study design limits ability to establish a causal relationship between hyperglycemia and DTG use. The absence of baseline plasma glucose measurements also hinders determination of the temporal relationship between hyperglycemia and DTG use.

The use of consecutive enrollment rather than random sampling and the limitation to participants attending the clinic on a specific day introduces selection bias, potentially affecting the representativeness of the sample.

Additionally, the study’s sample size was not adequately powered, which may have influenced the study results. The sample size calculation based on prevalence estimates from the adult population may not accurately reflect the prevalence of hyperglycemia in children and adolescents leading to an overestimation or underestimation of the required sample size, potentially affecting the precision and validity of the study findings.

The time period for the measurement of Random blood sugar after a meal was not standardized and therefore could have posted a falsely elevated plasma glucose level.

The study outcome which was hyperglycemia could have been over or under classified subsequently giving a relatively higher or lower prevalence respectively.

## Conclusions

The prevalence of Dolutegravir-related-hyperglycemia is high occurring in every 2 in 10 children and adolescents. The findings highlight the need for increased attention to glycemic screening and monitoring in this population. Health education talks on DTG adverse events were found to be protective against hyperglycemia underscoring the need to develop comprehensive health education messages that address DTG related complications and ensure optimal care for children and adolescents on Dolutegravir-based ART regimens.

## Recommendations

Based on the study findings, there should be advocacy for updates in HIV treatment guidelines to include specific recommendations for glycemic screening and monitoring for children and adolescents on Dolutegravir-based ART regimens.

The use of glycated hemoglobin (HbA1c) tests, for persons on a DTG based regimen should be supported given its accuracy in monitoring glycemic control over a longer duration.

Routine Glycemic screening and monitoring is recommended for children and adolescents on Dolutegravir-based ART regimens to support early detection and management of hyperglycemia.

Health Education programs for children, adolescents, and their caregivers should routinely focus on the potential side effects of Dolutegravir, including hyperglycemia with emphasis on the importance of lifestyle modifications, such as healthy eating habits and regular physical activity.

To conduct longitudinal studies to investigate the long-term implications of Dolutegravir-related hyperglycemia in children and adolescents while evaluating the progression of hyperglycemia, its impact on metabolic health, and the effectiveness of different management strategies.

## Data Availability

All relevant data are within the manuscript and its Supporting Information files.

## Acknowledgements

The authors would like to acknowledge the contribution of Ambrose Okibure for support in the statistical analysis and all the lecturers in the Department of Paediatrics and Child health, Makerere University for their insightful input from concept development to completion of the manuscript. We also appreciate Dr. Ben Watmon and Dr. Nathan Onyachi for providing clearance to conduct the study in Soroti and Lira RRHs.

